# Accumulation of Benzalkonium Chloride from Disinfectants in Dust Associated with Increased Microbial Tolerance

**DOI:** 10.64898/2026.04.14.26350823

**Authors:** Jiahe Yu, Shelby Tillema, Mary Akel, Abigail Aron, Estefania Espinosa, Stephanie A. Fisher, Tonia N. Branche, Leena B. Mithal, Erica M. Hartmann

## Abstract

Benzalkonium chloride (BAC) is widely used as a disinfectant in cleaning products and is frequently detected in indoor dust. In this study, we assessed dust samples, along with information on cleaning product use, from 24 pregnant participants. Dust samples were analyzed for BAC concentration and microbial tolerance. Different chain lengths of BAC (C12, C14, and C16) were quantified using LC–MS/MS, and bacterial isolates were tested for BAC tolerance using minimum inhibitory concentration (MIC) assays. BAC was ubiquitously detected, with C12 and C14 being dominant. Higher BAC concentrations were associated with reported disinfectant use and increased microbial tolerance. These findings suggest that indoor antimicrobial use may promote microbial resistance, highlighting potential exposure risks in indoor environments and the need for further investigation into health and ecological impacts.

## Introduction

Increased awareness of hand hygiene and infectious disease transmission has led to a rise in the use of disinfectants and cleaning products in indoor environments, making indoor environments the primary setting for antimicrobial chemical exposure. Especially following the 2016 ban of triclosan and several other chemicals and the subsequent COVID-19 pandemic, the use of quaternary ammonium disinfectants has increased dramatically. Quaternary ammonium compounds (QACs) are the most prevalent active ingredients in commonly used commercial and household products, as well as antimicrobial soaps. Among the 597 disinfectants listed by the U.S. Environmental Protection Agency (EPA) as effective against SARS-CoV-2 (List N), 277 products (46.3%) contain QACs.^1–3^ Concordant with their ubiquitous use, QACs have been detected across diverse environmental matrices, including wastewater sludge, surface water, sediments, soils, and dust.^4–8^ Exposure to QACs has been associated with adverse health effects.^9,10^ As the adverse health effects on humans are still being studied, gauging exposure levels in vulnerable groups is an essential first step to protect populations from adverse outcomes.

Among vulnerable populations, pregnancy and early infancy represent critical windows of susceptibility to both chemical and biological environmental exposures.^11^ Chemical exposures during these periods can disrupt developmental pathways, and due to smaller body size, higher concentrations per body weight in the fetus may be reached.^12^ Biological exposures are also especially influential *in utero* and during early life due to impact on the developing gut microbiota, which plays a critical role in development.^13^ Together, these exposures during early life can have long-lasting impacts on health.^12,14,15^ However, QACs exposure scenarios and exposure concentrations, as well as the associated risks related to altered microbial communities, have not been well characterized for pregnant individuals and infants in indoor environments. In this study, we focus on a major subgroup of QACs, Benzalkonium Chloride (BAC), and assess baseline exposure to pregnant individuals using data from a cohort study.

BAC is extensively used in disinfectant products, accounting for approximately 70% of total QACs.^1^ Different BAC chain lengths are present in different types of consumer products and consequently in different environments. In indoor dust, BAC with 12 and 14 carbon chain lengths (C12 and C14, respectively) are most abundant and represent more than 40% of all BAC.^16,17^ Indoor dust serves as a proxy for long-term indoor chemical exposure, reflecting the accumulation of BAC from routine cleaning product use. Concordant with the increased use of antimicrobial products during the COVID-19 pandemic, indoor dust concentrations of QACs increased markedly relative to pre-pandemic levels.^16^ The rapidly increasing environmental prevalence of BAC in indoor environments has generated public and scientific concern.

Beyond chemical exposures, antimicrobial products like BAC can exert selective pressure on indoor microbes, leading to altered microbial community function and increased resistance potential.^24^ Antimicrobial resistance (AMR) is one of the most critical global threats to public health, directly causing an estimated 1.27 million deaths and contributing to 4.95 million deaths worldwide in 2019.^25^ The spread of AMR is accelerated in part by the widespread use of antimicrobial chemicals and antibiotic medicines.^26^ Previous studies have shown that BAC can promote AMR in both natural and clinical environments,^27,28^ suggesting that similar selection pressures may occur in indoor microbial communities as well. However, the relationship between BAC concentrations in indoor residential environments and its influence on microbial tolerance remains largely unknown.

In this study, we analyzed indoor dust samples collected from 24 pregnant participants in the Chicago Perinatal Origins of Disease (CPOD) Study to investigate the potential for household BAC exposure and its impacts on indoor microbial communities. We quantified three common BAC chain lengths (C12, C14, and C16) in indoor dust using liquid chromatography–tandem mass spectrometry (LC–MS/MS) and assessed BAC tolerance in cultivable dust-associated bacteria using minimum inhibitory concentration (MIC) tests. We hypothesized that use of BAC-containing disinfectant products is associated with higher BAC concentrations in indoor dust, and elevated dust BAC concentrations are associated with increased BAC-tolerance in indoor microbes.

## Materials and Methods

### Sample and data collection

Household dust samples and cleaning product data were obtained from participants in the CPOD cohort, a prospective study that has enrolled pregnant individuals with a singleton gestation who attend prenatal care at two obstetric clinics in Chicago, Illinois since May 2023. All participants provided informed consent, and the study was approved by Institutional Review Boards of Northwestern and Lurie Children’s (IRB 2022-5510 and STU00218599, respectively) and was further approved by Erie Family Health’s Research Ethics Committee. Participants collected dust from inside their homes between 20 weeks 0 days and 36 weeks 6 days gestation using gloves and sterile tongs, typically from the collection bin of a household vacuum cleaner or from the manual collection of dust by sweeping. All samples were stored at −80 °C until analysis. Dust samples included in this analysis were collected from 06/2023 to 11/2023. At the time of dust sample collection, participants also self-completed an electronic questionnaire that ascertained the number of BAC-containing products used (e.g., brand name, ingredient list, etc.), type of products used (e.g., wipes, sprays, hand soap), frequency and purpose of use, and specific locations in the home where each product was applied. The full questionnaire is provided in the Supporting Information (Table S1).

### Sample processing

The approach used for chemical analysis in this study was adapted from Zheng et al.^16^ For each sample, at least 0.2 g of dust was weighed and transferred into a 15 mL centrifuge tube. Subsequently, 100 μL of internal standard and 2.5 mL of acetonitrile (99.7+%; Thermo Scientific Chemicals, Ward Hill, MA, USA) were added. The internal standard was prepared as a mixture of d7-BAC-C12, d7-BAC-C14, and d7-BAC-C16, each at a final concentration of 1 ppb. Then, samples were sonicated in a water bath for 1 hour and centrifuged at 2200x*g* for 15 minutes. After centrifugation, 1 mL of the supernatant was collected and transferred to a clean tube, to which 1 mL of acetonitrile (99.7+%; Thermo Scientific Chemicals, Ward Hill, MA, USA) was added. The extraction was repeated twice. The final combined extract volume was 3 mL. The pooled extract was filtered through a 0.45 μm PTFE syringe filter and analyzed at the Northwestern University Integrated Molecular Structure Education and Research Center (IMSERC).

### LC–MS analysis

An Agilent 6475 liquid chromatograph–triple quadrupole mass spectrometer was used to analyze the samples for targeted quantitative detection at IMSERC. The system was equipped with an Agilent Jet Stream electrospray ionization source operated in positive ion mode. Chromatographic separation was performed using an Agilent C18 column maintained at 40°C. The mobile phases consisted of (A) water containing 0.1% formic acid and (B) a 90:10 (v/v) isopropanol/acetonitrile mixture. The flow rate was set to 0.4 mL/min. The gradient program was as follows: 0–4 min, 50% A; 4–5 min, 20% A; 5–7 min, 0% A (100% B); and 7–7.1 min, 50% A. The system was then equilibrated at initial conditions. A 5 μL aliquot of each sample was injected into the LC system. MS data was acquired in multiple reaction monitoring mode for quantification of target analytes. All chemical solutions used for the analyses were HPLC grade or higher. The complete analyte list, details of instrumental parameters (Table S2), data analysis procedures, and representative chromatograms (Table S3) are provided in the Supporting Information.

### Sample enrichment, screening, and isolation of bacteria

The method used in this study was adapted from Fahimipour et al.^20^ For each sample, 20 mg of dust was weighed and suspended in 50 mL of sterile suspension buffer consisting of 42.5 mg/L KH_2_PO_4_ (Thermo Scientific Chemicals, MA), 250 mg/L MgSO_4_·7H_2_O (Thermo Scientific Chemicals, Ward Hill, MA, USA), 8.0 mg/L NaOH (Thermo Scientific Chemicals, Ward Hill, MA, USA), and 0.02% (v/v) Tween 80 (Thermo Scientific Chemicals, Ward Hill, MA, USA) in deionized water. The dust suspensions were placed on an orbital shaker, shaking at 180 rpm for 30 minutes. Following shaking, 100 µL of each suspension buffer was spread, in duplicate, onto Trypticase Soy Agar (Sigma-Aldrich, St. Louis, MO, USA) plates supplemented with 4 µg/mL itraconazole (TSA/I; 99%, Thermo Scientific Chemicals, Ward Hill, MA, USA) and incubated aerobically at 25°C for up to 6 days. After incubation, colonies were recorded and preliminarily identified based on morphological characteristics (Table S4).

To screen bacteria that can tolerate BAC, original TSA/I plates were replicated onto TSA/I with varying concentrations of BAC (95%+; MP Biomedicals, Irvine, CA, USA). Three BAC concentrations were used: 0 µg/mL as the positive control, 64 µg/mL for screening, and 1,000 µg/mL as the negative control. Replica plating was performed using sterile velveteen squares to transfer colonies from the master plate. All plates were incubated aerobically at 25°C for up to 6 days. BAC tolerance profiles were recorded for each isolate. Colonies growing on TSA/I containing 64 µg/mL BAC plates were selected as BAC-tolerant isolates. Colonies that did not grow on TSA/I containing 64 µg/mL BAC plates but were present on the original plates were classified as BAC-sensitive isolates. These isolates were cultured overnight in Tryptic Soy Broth (TSB), streaked twice onto TSA plates for purification, and stored at −80°C in 50% glycerol for further use.

### Antimicrobial tolerance assessment

Minimum inhibitory concentration (MIC) tests were performed on both BAC-tolerant isolates and representative BAC-sensitive isolates. An adapted methodology based on CLSI M100 and M07 was applied to guide the MIC test in this study.^29–31^ Each isolate was grown overnight in cation-adjusted Mueller–Hinton broth (CAMHB; Sigma-Aldrich, St. Louis, MO, USA) and subsequently inoculated into 96-well microtiter plates (Corning, NY, USA) at 50 µL per well. BAC working (95%+; MP Biomedicals, Irvine, CA, USA) solutions were prepared to generate concentration gradients specific to each isolated group. For BAC-tolerant isolates, the BAC gradient included 2000, 1024, 512, 256, 128, 64, and 0 µg/mL. For BAC-sensitive isolates, the gradient consisted of 512, 256, 128, 64, 32, 16, 8, 4, and 0 µg/mL. To each well, we added 50 µL of the corresponding BAC solution, resulting in a final volume of 100 µL. Six biological replicates were included for each concentration. Plates were incubated aerobically at 25°C for 24 hours, and optical density at 600nm (OD600) was measured at 24 hours to determine the MIC.

### Bacteria identification

Bacterial identification was performed using 16S rRNA gene Sanger sequencing. A subset of colonies exhibiting pronounced morphological differences was selected and subjected to colony PCR for taxonomic identification. The Q5 Hot Start High-Fidelity 2× Master Mix (New England Biolabs, Ipswich, MA) and universal 16S rRNA primers 27F and 1319R (Integrated DNA Technologies, Coralville, IA) were used to amplify the 16S rRNA gene region on a Mastercycler (Eppendorf, Hamburg, Germany) thermocycler. A 10-beta Competent *Escherichia coli* (New England Biolabs, Ipswich, MA, USA) and a blank reaction were included as positive and negative controls, respectively. PCR products were subsequently sent to ACGT (ACGT Inc., Wheeling, IL) for Sanger sequencing. The resulting sequences were processed and queried using BLASTn against the NCBI 16S rRNA sequences (Bacteria and Archaea) database (accessed Jan 25, 2026). to determine the closest taxonomic affiliations. Hits with E values ≤ 1×10^-16^ and high sequence similarity were considered reliable matches.

### Data analysis

Descriptive statistics for baseline participant characteristics, type and frequency of BAC-containing disinfectant products, the distribution of BAC concentration in household dust samples, and the distribution of MIC in BAC-sensitive and BAC-tolerant microbes in household dust samples are reported. Pearson correlation was performed to assess the correlation of 1) BAC concentrations with different chain lengths in indoor dust, 2) number of BAC-containing disinfectant products reportedly used by participants and total BAC concentrations in indoor dust, and 3) BAC concentrations in dust and the corresponding MIC of BAC-tolerant isolates based on MIC result. All statistical analyses and data visualization were conducted in R (v4.1.4) using packages including ggplot2 and Complex Heatmap.^32,33^ Statistical significance was defined as p ≤ 0.05, and p < 0.1 was considered indicative of a statistical trend.

## Results

We collected samples from a total of 31 participants, among which 27 paired dust samples were obtained and prioritized for analysis. After quality control, valid chemical analysis data were available for 24 participants. These 24 participants were therefore prioritized for subsequent analyses.

### Study population

Among 24 participants in the analytic cohort, 25% self-identified as non-White and 17% as Hispanic. Median age at enrollment was 33 years (IQR: 30-36) and median gestational age at dust collection was 19.4 weeks (IQR: 17-22).

### Reported disinfectant usage

Paired with the 24 dust samples included in this study, we received 24 completed survey responses including 80 photos of cleaning products. We cross-checked the survey responses with the photos provided to ensure the cleaning products were correctly identified and then used both the survey responses and photographic data to determine whether each product contained BAC. We assumed that participants were currently using all the cleaning products they reported in the questionnaire. Each BAC-containing product was counted separately, including products with identical ingredients used in different formats or locations. Participants reported using multiple BAC-containing disinfectant products (median: 3 products; range 0-5), and high frequency of cleaning household surfaces (median: 2 times per day; range 0-6). In total, 31.5% of the cleaning products contained BAC. Among all BAC-containing products, 39.4% were sprays and 60.6% were wipes. No BAC-containing soap usage was reported.

### BAC concentrations and composition in indoor dust

All 24 dust samples contained detectable levels of BACs. Total BAC concentrations (sum of C12, C14, and C16-BAC) ranged from 10.60 to 998.00 μg/g, with a median of 167.00 μg/g. Among individual chain lengths, C12-BAC was the most abundant, with concentrations ranging from 24.10 to 434.79 μg/g (median: 90.97 μg/g), followed by C14-BAC (1.09–490.51 μg/g; median: 52.09 μg/g) and C16-BAC (0.37–332.61 μg/g; median: 23.61 μg/g) (Figures 1 and 2). The median contribution of C12-, C14-, and C16-BAC to total BAC concentration in household dust were 57.7%, 27.0%, and 15.3%, respectively.

**Figure 1.**
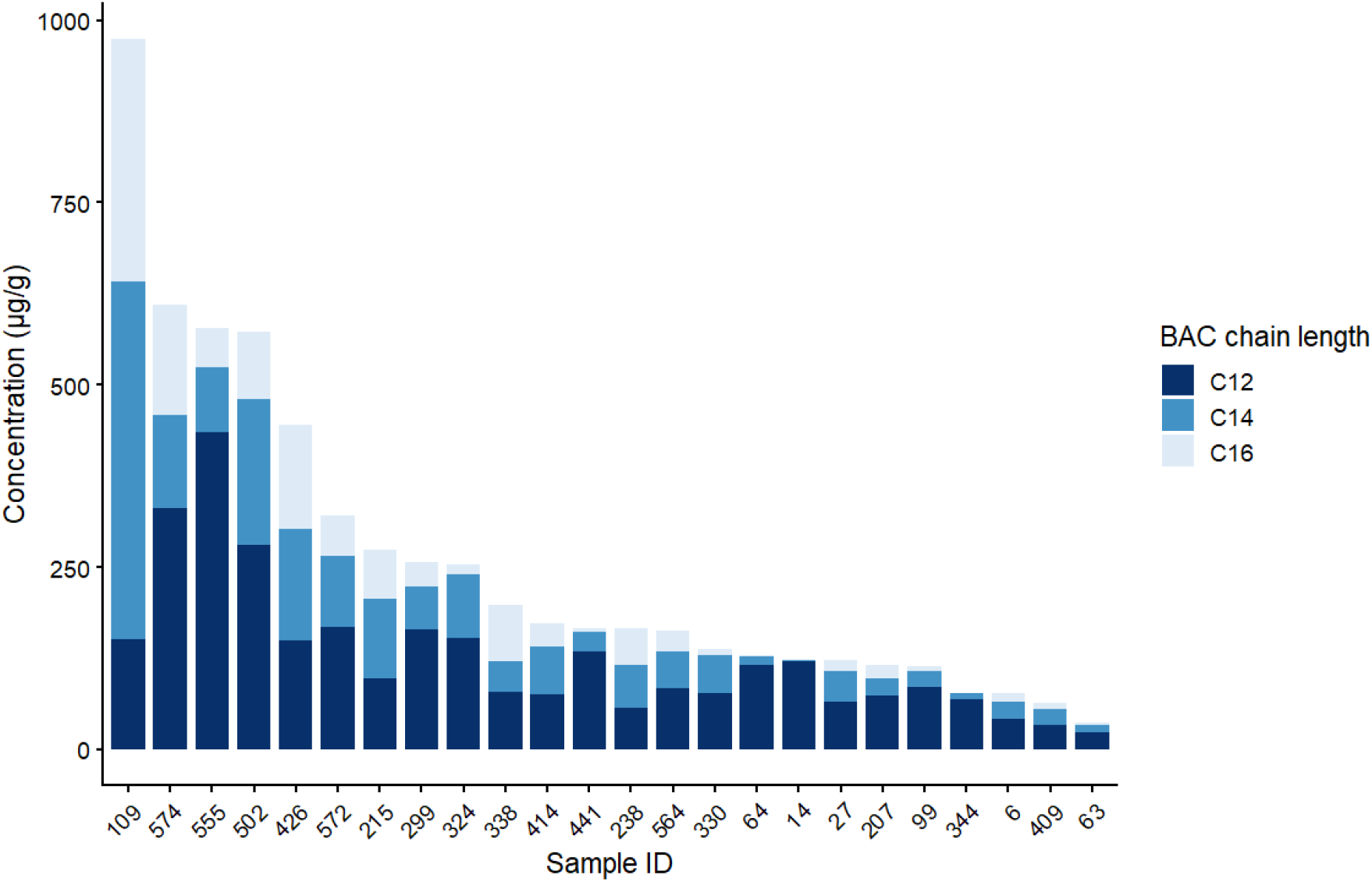
Concentrations and relative distributions of BAC chain length across 24 indoor dust samples, ordered from highest to lowest total BAC concentration. Individual bars represent the total BAC concentration in each sample, with color gradients indicating the contributions of C12, C14, and C16-BAC.

**Figure 2.**
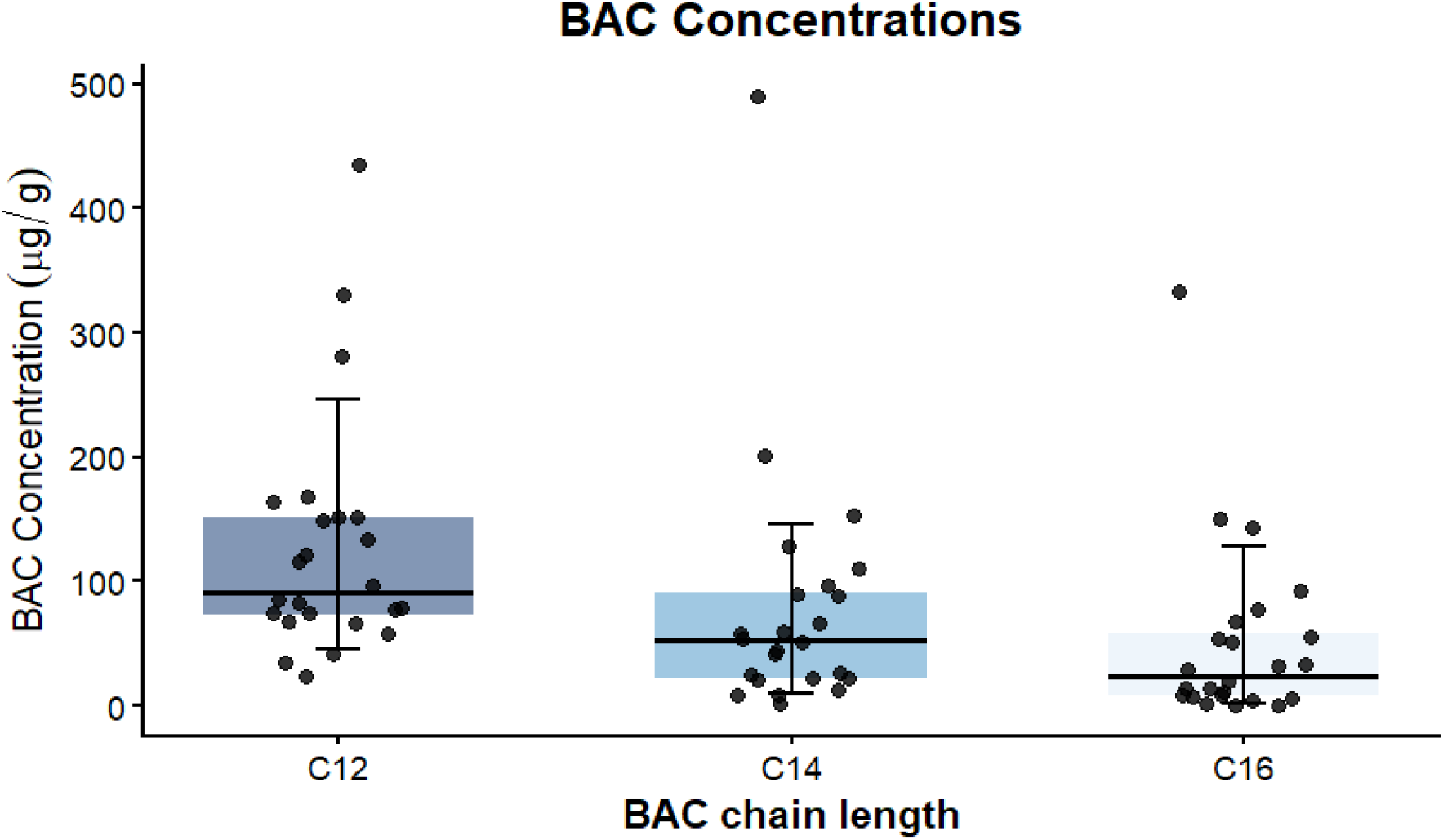
Distribution of BAC concentrations across different chain lengths. Individual points represent measured concentrations for each dust sample. The upper and lower horizontal lines indicate the 90th and 10th percentiles, respectively, while the colored bar denotes the 25th–75th percentile range. The central black line represents the median concentration for each BAC chain length.

**Figure 3.**
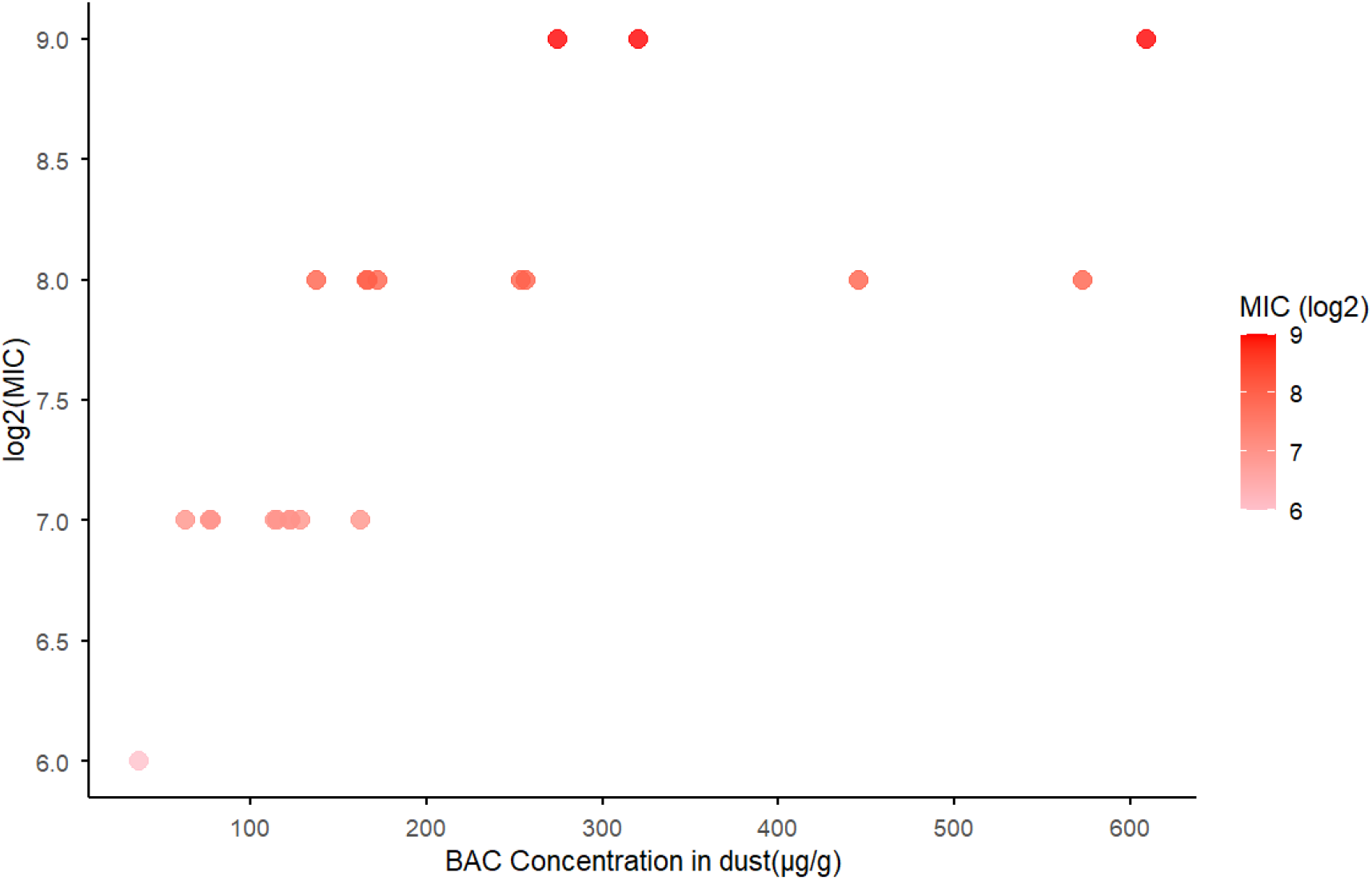
Relationship between BAC concentration in dust and log2-transformed MIC values for isolates collected from the dust. Points are colored by MIC. A significant positive correlation was observed (r = 0.711, p < 0.001).

Significant positive correlations were observed between C14-BAC and C16-BAC concentrations (r = 0.94, p < 0.05), whereas weaker correlations were observed between C12-BAC and C14-BAC (r = 0.35, p < 0.1) and between C12-BAC and C16-BAC (r = 0.36, p < 0.1). A positive correlation was observed between the number of BAC-containing disinfectant products reported and total BAC concentrations in indoor dust (r = 0.39, p < 0.05).

### Occurrence of BAC-tolerant bacteria in indoor dust microbial communities

Ten dust samples were first randomly selected and thoroughly mixed to generate an aggregate dust composite. Bacterial colonies displaying distinct morphologies, identified using a morphology characterization chart, were selected from the culture plates for MIC testing. The MIC results showed that only small subset (≈17%) of colonies could tolerate 64 μg/mL BAC (Table S5). Based on this preliminary result, 64 μg/mL BAC was subsequently applied as the selective concentration during screening. Morphological distinctions among colonies were documented, and the total number of distinct morphology types observed was recorded (Table S6).

Overall, we isolated 25 BAC-tolerant bacteria and 7 BAC-sensitive bacteria in the 24 dust samples. All isolates emerged within 24 hours of incubation at 25 °C and exhibited pronounced colony morphology differences. Of all colonies screened, 71.1% were classified as BAC-sensitive, indicating that most indoor dust bacteria remain susceptible to BAC disinfectants (Table S6). Only one dust sample did not yield any BAC-tolerant isolations. The BAC-sensitive isolates displayed minimal tolerance to BAC, with MIC values ranging from 0 to 8 μg/mL (median 4; IQR 0-4). In contrast, BAC-tolerant isolates showed substantially elevated MIC, ranging from 128 to 512 μg/mL (median 128; IQR 128-256). We further examined the relationship between the log-transformed BAC concentrations in dust and the corresponding MIC values of the recovered tolerant isolates and found a strong positive association (r = 0.711, p < 0.001).

Sanger sequencing results for selected colonies show that the isolates are predominantly affiliated with taxa commonly detected in indoor environments.^21^ The characteristics of these isolates, including MIC values, are summarized in Table 1. All original sequences provided at Supplementary Material.

**Table 1.**
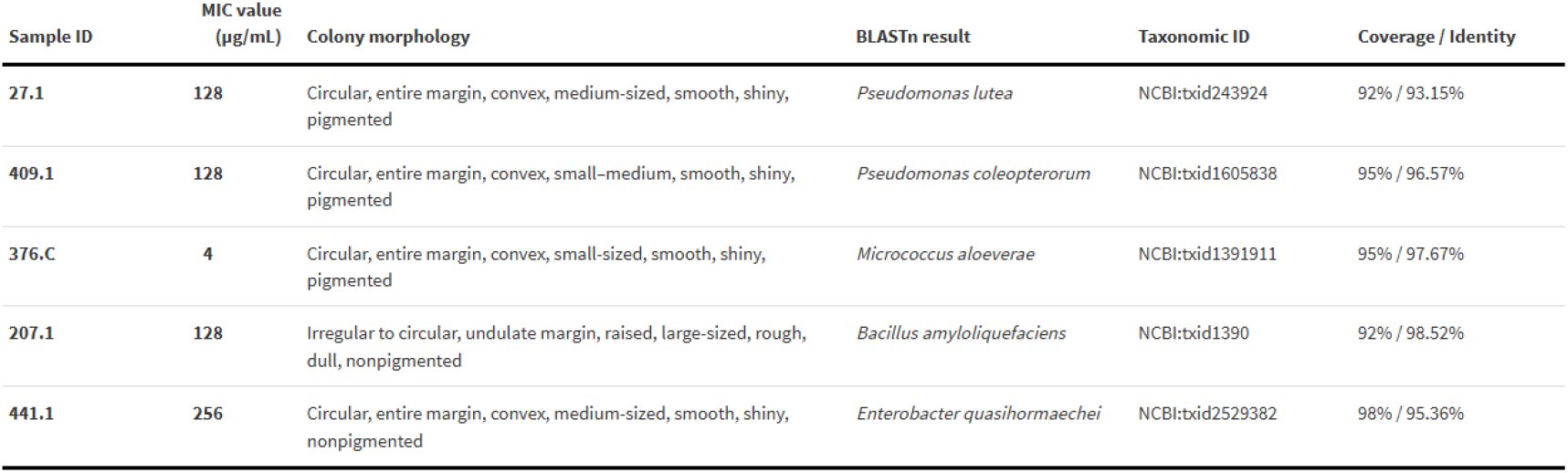
MIC values, colony morphology, and 16S rRNA BLASTn result of selected bacterial isolates. All colony morphology observations were made visually after 24 hours of growth. All results met threshold.

## Discussion

In this analysis of the CPOD cohort of pregnant participants and households in Chicago, we identified common use of BAC-based cleaning products that correlated with quantities of BACs in indoor household dust. Further, we identified BAC-tolerant bacterial organisms cultivated from dust, with higher MICs correlated with higher BAC concentrations in dust from that household indicating exposure-based tolerance. Our results corroborate previous studies demonstrating that the widespread use of BAC-based disinfectants in indoor environments leads to sub-inhibitory residual concentrations. We further provide evidence suggesting that use of BAC-based disinfectants promotes the emergence of BAC tolerance. By analyzing the use of BAC-containing disinfectants, their residual concentrations in indoor environments, and the responses of indoor microbial communities, this study provides novel insights for these fields.

In this analysis of the CPOD cohort, BAC containing disinfectant products are widespread across households of participants in Chicago. The compositional pattern we observed is consistent with previous reports of BAC distributions in post-COVID-19 indoor dust collected during the post-COVID-19 period samples.^16,17^ While median BAC concentrations in dust in our cohort are consistent with other studies, some CPOD households yielded dust samples with BAC concentrations an order of magnitude higher than the average, likely driven by differences in household practices.^16^

A significant positive correlation was observed between the use of multiple BAC-containing disinfectant products in households and BAC concentrations in indoor dust (Pearson correlation, r = 0.39, p < 0.05). However, no statistically significant correlation was observed between BAC concentrations in indoor dust and self-reported disinfection frequency from the questionnaire data (Pearson correlation; r = -0.04, p = 0.847). This may be attributable to variability in how participants defined and reported disinfection frequency, which could introduce uncertainty in the frequency estimates. Additionally, heterogeneity in the types and formulations of disinfectant products used across disinfection events may have further weakened this association.

The strong positive correlation between C14-BAC and C16-BAC suggests shared or closely related sources, whereas the weaker correlations involving C12-BAC indicate that this homolog may originate from more distinct usage patterns or product formulations. Moreover, the observed positive correlation between the number of BAC-containing disinfectant products reported and total BAC concentrations in indoor dust reinforces the role of household disinfectant use as a major contributor to indoor BAC accumulation. Together with the survey results, the correlation between BAC chain lengths indicate that the distribution of BAC in indoor dust is associated with the use of BAC-containing cleaning products.

Our microbiology results show that BAC tolerance is present within common indoor dust– associated bacteria and, consistent with previous results from other disinfectants, may be influenced by environmental disinfectant residues.^21^ Research on triclosan—an antimicrobial compound banned in 2016 due to health concerns—has demonstrated a significant positive correlation between its indoor concentrations and AMR.^20,34^ Given that BAC is commonly used as a substitute for triclosan, it may have a comparable influence on the development of tolerance. Our findings provide supporting evidence for this hypothesis by demonstrating an association between environmental BAC exposure and elevated bacterial tolerance. To compare our results against a clinical reference, two common clinically-relevant bacteria *Pseudomonas aeruginosa* and *Acinetobacter baumannii* typically exhibit BAC MIC values of 15-30 μg/mL and 10-50 μg/mL, respectively.^35–38^ The MIC we identified in BAC-tolerant bacteria was 128 to 512 μg/mL, suggesting that the use of BAC-containing disinfectants is associated with an increase in the highest tolerable BAC concentration of some bacteria. Nevertheless, the persistence of sensitive isolates indicates that selective antimicrobial pressure likely occurs prior to bacterial deposition in dust.

The identification of highly BAC-tolerant strains within ubiquitous environmental taxa suggests that tolerance is not confined to specialized microorganisms but can arise broadly within indoor microbiomes. Furthermore, the relationship between environmental BAC levels and bacterial tolerance supports the role of residual indoor disinfectants as a selective pressure shaping microbial communities. Collectively, these findings highlight the potential role of BAC-containing disinfectants in altering indoor microbial community composition by promoting the persistence of tolerant subpopulations.

Together, our data suggest that BAC exposure in indoor environments is ubiquitous and increases with the usage of more BAC-containing cleaning products. Our results further suggest that BAC usage alters the surrounding indoor microbial community. Especially for vulnerable populations, increased BAC exposure and exposure to a more antimicrobial tolerant microbial community may have important health impacts later in life.

### Implications and Limitations

This study has several limitations. First, data on disinfectant use were self-reported and may be subject to recall bias or inaccurate description, although cross-validation with product photographs did mitigate some potential misclassification. Second, our analysis focused only on three common BAC chain lengths and did not examine other QAC compounds that may also impact the indoor chemical and microbial environment. Our observations are correlative and do not establish a causal relationship between the presence of BAC in dust and the emergence of BAC-tolerant bacteria. In addition, dust samples were frozen prior to processing, potentially biasing the recovered community.

Further, our MIC analysis was performed under a single culture condition, which likely captured only a subset of the antimicrobial-resistant bacteria present in the dust, excluding organisms that were unable to grow under the chosen conditions. Moreover, the molecular mechanisms underlying BAC resistance remain unclear. Although previous studies suggest that efflux pumps are a primary mechanism for microbial resistance to BACs, further experiments are needed to validate these findings in the context of indoor dust microbial communities.^27,39^

Despite these limitations, this study fills a critical knowledge gap by characterizing linked disinfectant use data with environmental residue measurements and microbial response within residential indoor environments among a vulnerable population of pregnant people, with potential implications for fetal development and the postnatal microbiome. Our findings from a well-characterized prospective cohort suggest that widespread BAC use may create selective conditions favoring increased microbial tolerance, which could represent an early stage in the development of antimicrobial resistance. Understanding these impacts is critical, as the use of BAC-containing disinfectants is rapidly increasing. This study provides important guidance for the management of BAC exposure and the mitigation of AMR, and their unique interaction in indoor environments.

## Supporting information

Sanger sequencing data

Full survey

## Data Availability

Source code for the analysis is available: https://github.com/Reaction1337/BAC_AMR_Indoor_Coding.

Bacteria isolates are available upon request.

## Supporting Information

For chemical analysis, the chemical and reagents, analytical instrument, quality assurance and quality control, mass spectrometry parameters and the representative chromatograms of the internal standard are provided in Supporting Information.

For participants data, a survey template is provided in Support Information.

## Acknowledgement

This study was partially funded by The Founders’ Board of Ann and Robert H. Lurie Children’s Hospital of Chicago. Research reported in this publication was also supported, in part, by the National Institutes of Health’s National Center for Advancing Translational Sciences, Grant Number UM1TR005121.

This work made use of the IMSERC (RRID:SCR_017874) MS facility at Northwestern University, which has received support from the Soft and Hybrid Nanotechnology Experimental (SHyNE) Resource (NSF ECCS-2025633), the State of Illinois, and the International Institute for Nanotechnology (IIN).

We are deeply grateful to the members of the Hartmann group for their support and assistance with this research and troubleshooting.

We gratefully acknowledge Steve Klimcak and Dr. Fernando ‘Ralph’ Tobias for their invaluable contributions to the LC-MS analysis.

