## Supplementary Information for "Accumulation of Benzalkonium Chloride from Disinfectants in Dust Associated with Increased Microbial Tolerance"

Support Information for Accumulation of Benzalkonium Chloride from Disinfectants in Dust Associated with Increased Microbial Tolerance

**Jiahe Yu, BS[1]**; Shelby Tillema, MS[1]; Mary Akel, MPH[2]; Abigail Aron, BA[2]; Estefania Espinosa, BA[2]; Stephanie A. Fisher, MD, MPH[2]; Tonia N. Branche, MD, MPH[3]; Leena B. Mithal, MD, MSCI[2]; Erica M. Hartmann, PhD[12*]

[1]Department of Civil and Environmental Engineering, Northwestern University, Evanston, Illinois, USA

[2]Feinberg School of Medicine, Northwestern University, Chicago, Illinois, USA

[3]Ann & Robert H. Lurie Children's Hospital of Chicago, Chicago, IL, USA

*Chemical and reagents.* Three native standards, including Zephirol-d7(C12-BAC), Benzyldimethyltetradecylammonium-d7 Chloride(C14-BAC) and Cetalkonium Chloride-d7(C16-BAC) were all obtained from Toronto Research Chemicals. All solvents and chemicals used in this study were HPLC grade or higher.

*Analytical Instrument.* An Agilent 1290 Infinity II Ultra-high Performance Liquid Chromatograph system coupled with Agilent 6475 Triple-quadrupole mass spectrometer (Agilent 1290 Infinity II UHPLC – 6475 QQQ-MS) was used for the analysis in this study. The instrument was provided by Integrated Molecular Structure Education and Research Center (IMSERC), a core facility at Northwestern University. The UHPLC analysis used an Anilgent UHPLC InfinityLab Poroshell 120 EC-C18 column, Inner Diameter (ID) 2.1mm, Particle Size 2.7 µm. Nitrogen gas was used as nebulizer. The nebulizer pressure, gas flow, gas temperature, capillary voltage, sheath gas temperature, and sheath gas flow were set to 40 psi, 10 L/min, 400 °C, 3500 V, 400 °C, and 12 L/min, respectively.

*Quality assurance and quality control.* Six blank solvents samples and two internal standard samples are obtained each run with the dust samples. Blank samples constituted less than 0.1% of the sample levels. Method detection limits (MDLs) were based on a signal to-noise ratio of three. The MDLs were as follows: BAC-C12, 0.0002 mg/L; BAC-C14, 0.0001 mg/L; BAC-C16, 0.0001 mg/L. Fifteen dust samples were randomly selected and combined to create aggregate dust for quality control. The aggregate dust was used in a spike-recovery test, with each analyte spiked at 100 ng. The mean recoveries (± standard error) were as follows: BAC-C12, 106.1 % ± 7.4 %; BAC-C14, 154.5 % ± 10.8 %; and BAC-C16, 157.3 % ± 7.1 %. The spike recovery rate was above 100%, which may be related to background concentration and matrix interference but are all acceptable ranges.

*Data Analysis.* All data analyses were performed on the IMSERC platform, and chemical quantification was conducted using QQQ-MS instrumentation.

**Table S1.** Template for participant survey

### Cleaning Practices Questionnaire (~5-10 minutes)

These questions ask about your household cleaning routines and products you use, so we can better understand the chemical environment of your home.

**Household Cleaning Practices (Prácticas de limpieza del hogar) Please answer the following questions regarding your home environment and the types of cleaning products you use.**

**Responda las siguientes preguntas sobre el entorno de su hogar y los tipos de productos de limpieza que utiliza.**

What are the top 3-5 cleaning products you use at home to clean your surfaces and bathrooms (names/brands)?

¿Cuáles son los principales 3 a 5 productos de limpieza que usa en casa para limpiar las superficies y baños (nombres/marcas de productos)?

________________________________________

Could you upload a picture of the product label(s)?

¿Puede subir fotos de las etiquetas de los productos?

Feel free to upload a picture of the products as a group. Please also make sure the product label is visible in the photo. Puede cargar una imagen de los productos como un grupo. Asegúrese también de que la etiqueta del producto esté visible en la foto.

What products do you specifically use to clean the surfaces in your baby's room or your baby's items?

¿Qué productos utiliza específicamente para limpiar las superficies de la habitación de su bebé o los artículos de su bebé?

________________________________________

Could you upload a picture of the product label(s)?

¿Puede subir fotos de las etiquetas de los productos?

Feel free to upload a picture of the products as a group. Please also make sure the product label is visible in the photo. Puede cargar una imagen de los productos como un grupo. Asegúrese también de que la etiqueta del producto esté visible en la foto.

For full survey, please check Cleaning Practices Questionnaire in supporting information files.

**Table S2.** Mass spectrometry parameters

| *Compound* | *Retention time (min)* | *Precursor ion* | *Fragmentor (volts)* | *Product ions (m/z)* | *Collision energy (volts)* |
|---|---|---|---|---|---|

| | | | | | |
|---|---|---|---|---|---|
| C12-BAC | 1.17 | 304.3 | 120 | 91.0 | 41 |
| | | | | 212.2 | 25 |
| C14-BAC | 1.74 | 332.4 | 110 | 91.0 | 41 |
| | | | | 240.3 | 25 |
| C16-BAC | 2.89 | 360.5 | 91 | 91.0 | 41 |
| | | | | 268.2 | 25 |

**Table S3.** The representative chromatograms of the internal standard(1ppb)

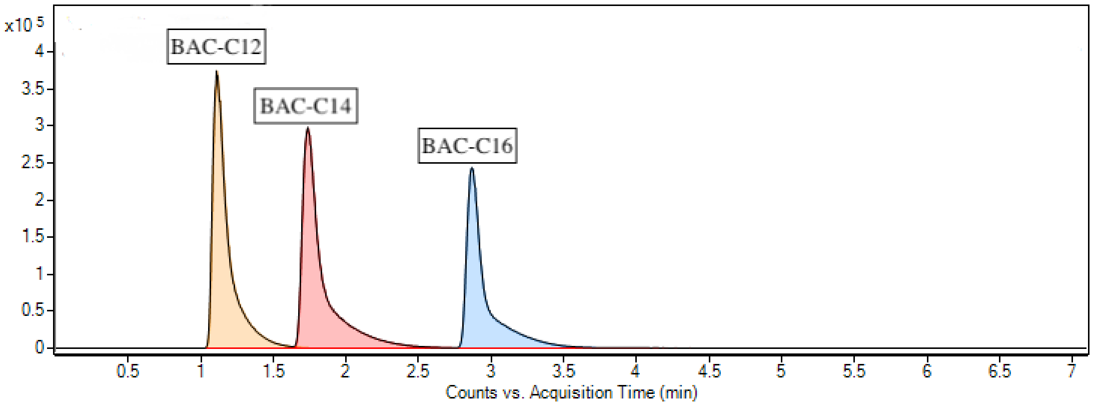


**Table S4.** Morphology characterization chart

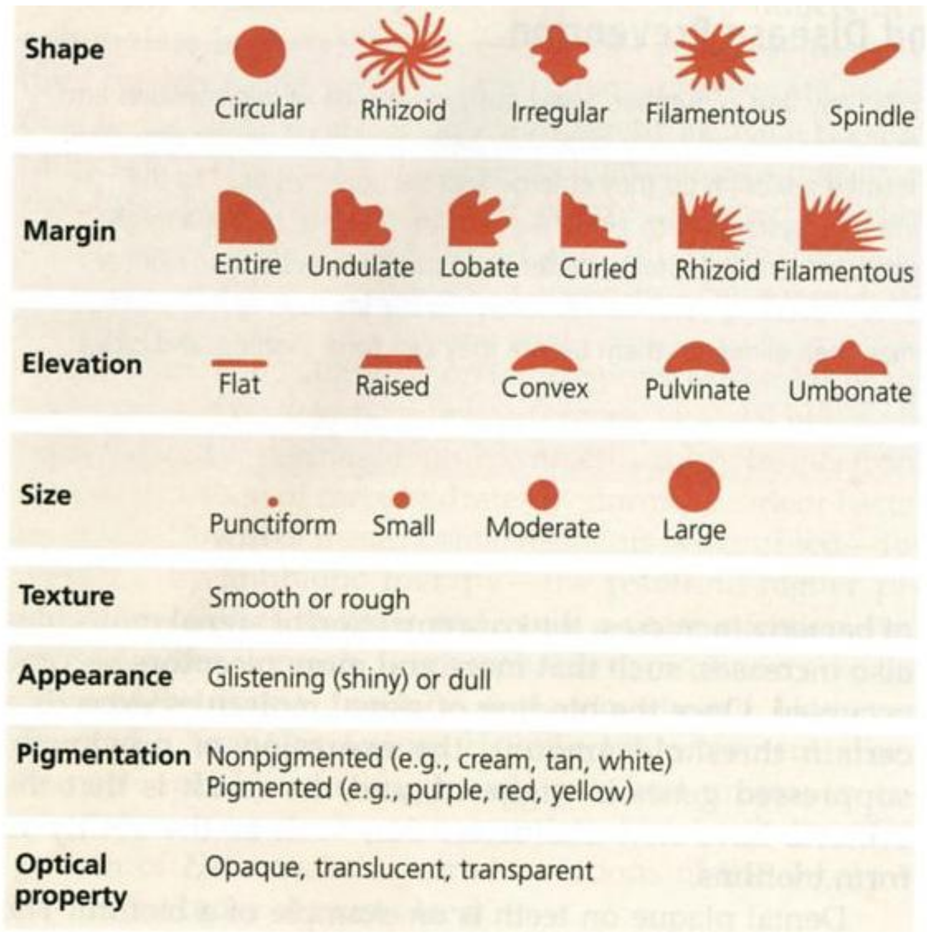

**Table S5.** Aggregate dust culture colonies MIC result

| *Colony ID* | *6hr mic* | *12hr mic* | *24hr mic* |
|---|---|---|---|
| Agg.1 | 4 | 4 | 4 |
| Agg.2 | 128 | 64 | 64 |
| Agg.3 | 8 | 4 | 4 |
| Agg.4 | 4 | 4 | 4 |
| Agg.5 | 16 | 8 | 8 |
| Agg.6 | 32 | 32 | 32 |

**Table S6.** Colonies show different types of morphology between TSA/I plate and 64 μg/mL BAC TSA/I plate.

| Sample ID | TSA/I plate | 64 μg/mL BAC TSA/I plate |
|---|---|---|
| 64 | 5 | 2 |
| 324 | 3 | 1 |
| 99 | 8 | 2 |
| 330 | 6 | 2 |
| 344 | 7 | 1 |
| 81 | 3 | 1 |
| 109 | 4 | 1 |
| 14 | 2 | 1 |
| 502 | 2 | 1 |
| 564 | 3 | 1 |
| 555 | 2 | 1 |
| 6 | 4 | 1 |
| 426 | 3 | 1 |
| 212 | 2 | 1 |
| 299 | 2 | 1 |
| 27 | 3 | 1 |
| 338 | 2 | 1 |
| 441 | 4 | 1 |
| 409 | 3 | 1 |
| 574 | 3 | 1 |
| 215 | 4 | 1 |
| 376 | 4 | 1 |
| 207 | 4 | 0 |
