## Supplementary material for "Accumulation of Benzalkonium Chloride from Disinfectants in Dust Associated with Increased Microbial Tolerance": Full survey

### Cleaning Practices Questionnaire (~5-10 minutes)

These questions ask about your household cleaning routines and products you use, so we can better understand the chemical environment of your home.

**Household Cleaning Practices (Prácticas de limpieza del hogar) Please answer the following questions regarding your home environment and the types of cleaning products you use.**

**Responda las siguientes preguntas sobre el entorno de su hogar y los tipos de productos de limpieza que utiliza.**

What are the top 3-5 cleaning products you use at home to clean your surfaces and bathrooms (names/brands)?

¿Cuáles son los principales 3 a 5 productos de limpieza que usa en casa para limpiar las superficies y baños (nombres/marcas de productos)?

__________________________________

Could you upload a picture of the product label(s)?

¿Puede subir fotos de las etiquetas de los productos?

Feel free to upload a picture of the products as a group. Please also make sure the product label is visible in the photo. Puede cargar una imagen de los productos como un grupo. Asegúrese también de que la etiqueta del producto esté visible en la foto.

What products do you specifically use to clean the surfaces in your baby's room or your baby's items?

¿Qué productos utiliza específicamente para limpiar las superficies de la habitación de su bebé o los artículos de su bebé?

__________________________________

Could you upload a picture of the product label(s)?

¿Puede subir fotos de las etiquetas de los productos?

Feel free to upload a picture of the products as a group. Please also make sure the product label is visible in the photo. Puede cargar una imagen de los productos como un grupo. Asegúrese también de que la etiqueta del producto esté visible en la foto.

What hand soap do you use for yourself and your baby?

¿Qué jabón de manos usa para usted y su bebé?

__________________________________

Could you upload a picture of the product label(s)?

¿Puede subir fotos de las etiquetas de los productos?

Feel free to upload a picture of the products as a group. Please also make sure the product label is visible in the photo. Puede cargar una imagen de los productos como un grupo. Asegúrese también de que la etiqueta del producto esté visible en la foto.

---

What kind of cleaning wipes do you use in your household?

¿Qué tipo de toallitas de limpieza utiliza?

__________________________________

---

How often do you use these wipes?

¿Con qué frecuencia utiliza estas toallitas?

- ◯ Daily/diariamente
- ◯ Weekly/semanalmente
- ◯ Every two weeks/cada dos semanas
- ◯ Monthly/mensualmente
- ◯ Less often/con menos frecuencia
- ◯ Never use cleaning wipes/Nunca use toallitas limpiadoras

---

Do any of your cleaning products say "antimicrobial," "antibacterial," or "disinfectant"?

¿Alguno de sus productos de limpieza dice "antimicrobiano", "antibacteriano", o "desinfectante"? ¿Usa desinfectantes para limpiar su casa?

- ◯ Yes
- ◯ No

---

What products are these? (name(s)/brand(s))

¿qué producto?

__________________________________

---

How often do you use these products?

¿con cuánta frecuencia usa estos productos?

- ◯ Daily/diariamente
- ◯ Weekly/semanalmente
- ◯ Every two weeks/cada dos semanas
- ◯ Monthly/mensualmente
- ◯ Less often/con menos frecuencia

---

Do you have pets that live in the home?

¿Tiene mascotas que viven dentro de la casa?

- ◯ Yes
- ◯ No

---

How many?

¿cuántas?

__________________________

---

What kind of pet(s) do you have?

¿qué tipo de mascotas tiene?

__________________________________

How long have you had your pets?

¿por cuánto tiempo ha tenido las mascotas?

Please report the length of time in years. If you've had your pets for less than 1 year, roud up to 1 year.

Por favor informe la duración del tiempo en años. Si ha tenido a sus mascotas por menos de 1 año, redondee hasta 1 año.

________________________________

If other, please describe

Si es otro, por favor describa

________________________________

What year was your house/building built?

¿En qué año se construyó su casa/edificio?

If you don't know, please enter "don't know".

Si no lo sabe, ingrese "no lo sé".

________________________________

Do you have a yard or outdoor space?

¿Tiene jardín o un espacio al aire libre?

- ◯ Yes
- ◯ No

What size is your home in square feet? (an estimation is ok)

¿De qué tamaño es su casa (en pies cuadrados)?

________________________________

What kinds of flooring do you have in your home?

¿Qué tipo de suelo tiene en su casa?

Select all that apply.

Seleccione todas las que correspondan.

- ☐ Carpet/Alfombras
- ☐ Tile/baldosas (losetas)
- ☐ Hardwood/madera dura
- ☐ Vinyl/Laminate/pisos de vinilo
- ☐ Concrete/concreto
- ☐ Rugs/alfombras
- ☐ Other/Otro

If other, please describe

Si es otro, por favor describa

________________________________

What kind of cooling/heating does your home have?

¿Qué tipo de refrigeración/calefacción tiene su casa?

Select all that apply.

Seleccione todas las que correspondan.

- ☐ Central heat/air/Aire central
- ☐ Radiator heat/calor de radiador
- ☐ Window AC unit/unidad de aire acondicionado de ventana
- ☐ Wood stove/estufa de leña
- ☐ Gas stove/estufa de gas (para calefacción)
- ☐ Other/Otro
- ☐ No heating or cooling systems/sin sistemas de calefacción o refrigeración

If other, please describe

Si es otro, por favor describa

________________________________

| | |
|---|---|
| How often do you use this appliance(s)?<br><br>¿Con qué frecuencia utiliza este aparato? | ◯ Daily/diariamente<br>◯ Weekly/semanalmente<br>◯ Every two weeks/cada dos semanas<br>◯ Monthly/mensualmente<br>◯ Less often/con menos frecuencia |
| Do you ever open your windows?<br><br>¿Alguna vez abre las ventanas? | ◯ Yes<br>◯ No |
| How often do you do this?<br><br>¿Con qué frecuencia? | ◯ Daily/diariamente<br>◯ Weekly/semanalmente<br>◯ Every two weeks/cada dos semanas<br>◯ Monthly/mensualmente<br>◯ Less often/con menos frecuencia |
| Do you have an air filtration system or air purifier?<br><br>¿Tiene un sistema de filtración de aire o purificador de aire? | ◯ Yes<br>◯ No |
| How often do you use it?<br><br>¿con cuánta frecuencia lo usa? | ◯ Daily/diariamente<br>◯ Weekly/semanalmente<br>◯ Every two weeks/cada dos semanas<br>◯ Monthly/mensualmente<br>◯ Less often/con menos frecuencia |
| Do you have a humidifier or de-humidifier?<br><br>¿Tiene un humidificador o deshumidificador? | ◯ Yes<br>◯ No |
| How often do you use it?<br><br>¿con cuánta frecuencia lo usa? | ◯ Daily/diariamente<br>◯ Weekly/semanalmente<br>◯ Every two weeks/cada dos semanas<br>◯ Monthly/mensualmente<br>◯ Less often/con menos frecuencia |
| What is your primary source of drinking water?<br><br>¿Cuál es su principal fuente de agua potable? | ☐ Tap water/agua del grifo<br>☐ Filtered water (through fridge or filtered dispenser)/Agua filtrada (a través del refrigerador o dispensador filtrado)<br>☐ Bottled water/Agua embotellada<br>☐ Other/Otro |
| If other, please describe<br><br>Si es otro, por favor describa | ________________________ |
| What is the average temperature in your home currently? (in farenheit)<br><br>¿Cuál es la temperatura promedio en su hogar actualmente? en farenheit | ____________________ |
| What is the average humidity in your home currently? (in percent)<br><br>¿Cuál es el nivel de humedad promedio en su hogar actualmente? (en porcentaje) | ____________________ |

| | |
|---|---|
| Is there any visible or significant water damage in your home that you know of?<br><br>¿Hay algún daño de agua visible o significativo en su hogar? | ◯ Yes<br>◯ No |
| When you collected your dust sample, did you collect from a vacuum or using another method (sweeping/swifter/etc.)? Select all that apply<br><br>Cuando colecto su muestra de polvo, ¿la recogió con una aspiradora o utilizó otro método (barrido/rápido/etc.)? Seleccione todas las que correspondan | ☐ Vacuum (aspiradora)<br>☐ Other method (otro metodo) |
| From which rooms did you collect dust when you submitted environmental samples? (select all that apply)<br><br>¿En qué habitaciones recogió polvo cuando envió muestras ambientales? (seleccione todas las que correspondan) | ☐ Bedrooms (Cuartos)<br>☐ Living room (Sala)<br>☐ Bathrooms (Baño)<br>☐ Kitchen (Cocina)<br>☐ Outdoor area (garage or patio)<br>☐ Whole house (toda la casa)<br>☐ Other (otro) |
| If other, please describe: | __________________________________ |
